# An SEIDR Model for the Early Spread of COVID-19

**DOI:** 10.1101/2023.02.17.23286115

**Authors:** Tingrui Cao

## Abstract

In this article, we conduct a literature review on the history and mathematical modeling of infectious diseases and COVID-19. Next, some simple epidemic dynamic models and the basic reproductive number theory are introduced. We propose a SEIDR model for COVID-19 and provide the solution methods for the basic reproduction number, parameters, and dynamic model. Finally, we simulate the early stages of the COVID-19 epidemic in Argentina, Indonesia, Mexico, and South Africa with the SEIDR model.

## 1 Background

In this part, we will first introduce the history of infectious diseases and specifically introduce the origin and development of COVID-19 in Section 1.1. This will be followed by an introduction to the history of mathematical modeling of infectious diseases, with an introduction to the research on mathematical modeling of COVID-19 in Section 1.2.

### 1.1 Epidemics and COVID-19

Infectious disease (epidemic) is a disease caused by pathogens (including viruses, bacteria, fungi, parasites, etc.) that can spread between humans or between humans and animals and spread rapidly. Throughout human development, the prevalence of infectious diseases has never stopped. Every occurrence brings massive disasters to humanity and even affects the development track of human history. Well-known infectious diseases, including plague (Black Death), smallpox, cholera, tuberculosis, malaria, Ebola, dengue fever, SARS, etc., have all caused great harm to human beings. In 430 BC, a plague broke out in the ancient Greek city of Athens and lasted for three years. Nearly half of the city’s population died, and the Athenian civilization declined [1]. Smallpox that broke out in the ancient Roman Empire between 165 and 180 AD caused as many as 25% population [2]. In the 14th century, a large-scale plague broke out in Europe, killing 25 million people. By the end of World War I, the Spanish flu had infected 1 billion people worldwide. In 2003, the SARS virus ravaged China [3]. In 2009, the H1N1 flu claimed tens of thousands of lives in the United States [4]. From 2012 to 2015, Middle East Respiratory Syndrome (MERS) caused tens of thousands of infections [5]. In the more than 100 years from the last century to the present, the total number of deaths caused by infectious diseases has reached 1.68 billion, which is more than ten times the number of deaths in wars [6]. In addition, a series of problems such as social disorder, economic regression, and famine caused by infectious diseases has also seriously affected the development of human society.

In December 2019, a respiratory infectious disease caused by a novel coronavirus was first discovered in Wuhan, China, and spread rapidly in Wuhan [7]. On February 11, 2020, the International Committee on Taxonomy of Viruses (ICTV) officially issued a statement, naming the virus SARS-CoV-2, and the disease caused by the virus was named COVID-19 by the World Health Organization (WHO) [8]. Since COVID-19 has spread worldwide, e.g. [9] and [10]. Compared with the SARS virus that broke out in China in 2003, the incubation period of COVID-19 is significantly longer. An early study found that the average incubation period for COVID-19 was 5.2 days [11]. Later results showed that its average incubation period might be 3.0 days [12] or 4.75 days [13]. It is worth noting that there are a certain number of asymptomatic infections of COVID-19 [10], and its fatality rate is much lower than that of SARS, and MERS [12]. As of January 29, 2020, the cumulative number of confirmed cases of COVID-19 has surpassed that of SARS. Uncertainty in the incubation period, contagious asymptomatic infections, and the super-transmissibility of the virus has brought great challenges to the control of the epidemic. The outbreak of COVID-19 not only seriously endangers the lives and health of people worldwide but also undermines society’s stability. The economies of various countries have been greatly impacted, and many workers have been forced to lose their jobs.

### 1.2 Research of Epidemic in Mathematics

In the long history of human struggle against infectious diseases, mathematicians have been trying to use mathematical models to study the law of the spread of infectious diseases. Mathematical models can explore the changes in conditions in different situations and predict the development trend of diseases. As early as 1760, Bernoulli used mathematical tools to study the transmission characteristics of smallpox, starting the research of mathematics in epidemiology [14]. In 1906, Harmer established a mathematical model of a discrete-time system to study the recurrence of measles and, for the first time, proposed the concept of morbidity, that is, the frequency of new occurrence of a particular disease in a specific population within a certain period [15]. In 1926, Kermack and McKendrick proposed the landmark SIR compartment model [16] of epidemiology through the study of the Black Death that broke out in London in the 17th century and the plague that broke out in Mumbai in the 20th century. Then the two scholars proposed the SIS compartment model [17] in 1932 and obtained the “threshold theory” which pointed out the relationship between the prevalence of infectious diseases and the basic reproductive number, which laid the foundation for the vigorous development of infectious disease dynamics in the future. Since then, more infectious disease models have evolved based on the idea of the warehouse model, such as SIRS [18], SEIR [19], etc. Since the 1950s, scholars have established various infectious disease models, including stochastic differential equation model [20], timedelay integral-differential equation model [21], impulse differential equation model [22], partial differential equation model [23], etc. to describe the spread of infectious diseases. At the same time, computer technology and statistical analysis methods are applied to infectious disease models. In 2006, Lekone et al. [24] applied the SEIR model to simulate the spread of the Ebola virus and used the maximum likelihood estimation and Monte Carlo Markov Chain (MCMC) method to estimate the parameters in the model. In 2014, based on the Ebola epidemic in West Africa, Ellina et al. [25] used the SEIR model to fit the infection data of Guinea, Sierra Leone, and Liberia and sought the optimal control strategy from the perspective of optimization.

Since the outbreak of COVID-19, mathematicians never stop studying the disease. In the early stage, the estimation of the reproduction number and final size of COVID-19 are given [26, 27, 28]. In [29], Mehra et al. built an SQAIR compartment to simulate the epidemic in Korea. Public behavior and government action as two important control factors have been analyzed by a nonlinear dynamic model in [30]. In [31], the physics-informed neural network is utilized to solve integer and fractional order COVID-19 epidemiological models.

## 2 Epidemic Dynamic Models

Analyzing and describing the spread of epidemics through models is an essential method in the dynamics of epidemics. Establishing a proper mathematical model can reflect the characteristics of the disease based on the disease transmission mechanism, disease characteristics, biological factors, and social control. Through the qualitative and quantitative analysis of the model and numerical fitting and simulation combined with actual data, the epidemic law of the epidemics can be revealed, the law of its development and change can be predicted, the critical factors of the spread of the infectious disease can be analyzed, and the best way to control and prevent the disease can be found. These models have enabled people to have a deeper understanding of the laws of the spread of epidemics. The basic reproductive number is an important index to evaluate the transmission ability of epidemics. We first introduce two classic epidemic dynamic models–the SIR model in Section 2.1 and the SEIR model in Section 2.2. Then the basic reproductive number theory is presented in Section 2.3.

### 2.1 SIR Model

In this section, we first introduce the classic SIR model in Section 2.1.1. Based on this, a SIR model with birth and death is presented in Section 2.1.2.

#### 2.1.1 Classic SIR Model

The classic SIR model was proposed in [16]. This model describes an epidemic with the following characteristics: the infected person can recover after some time, and after becoming a recovered person, he is immune to the disease and will not be reinfected with the disease for life. For example, measles and chickenpox are suitable for the SIR model. Based on this, we divided the population into three groups: susceptible (*S*), infected (*I*), and recovered (*R*). The numbers of these three groups of people at time *t* are respectively represented by *S*(*t*), *I*(*t*), and *R*(*t*). The classic SIR model considers short-term epidemics, so birth and death factors are not considered. The flowchart of the classic SIR model shows in Figure 1, where *β* is the contact rate and *γ* is the recovery rate.

**Figure 1:**
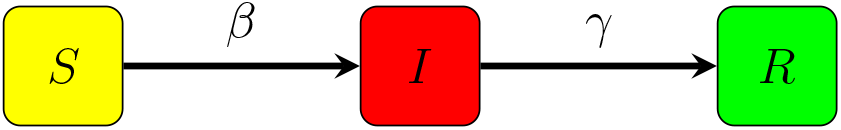
Flowchart of classic SIR model

Based on the assumptions and flowchart above, the mathematical form of the SIR model is presented as (2.1).

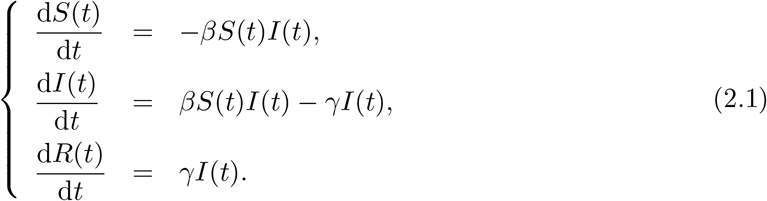

#### 2.1.2 SIR Model with Birth and Death

In this section, we consider a class of daily epidemics, which last for a long time, so it is necessary to consider births and deaths within the time of disease onset. The flowchart of the SIR model with birth and death is given as Figure 2, where *c* is the natural fatality rate, *d* is the epidemic fatality rate, and *μ* is the natural birth rate. Assuming the total population is *N*, we have:

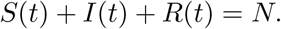

**Figure 2:**
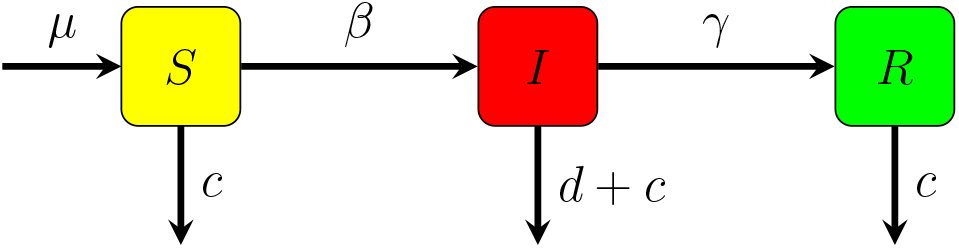
Flowchart of SIR model with birth and death

Under the assumption that *μ* > *c* and *d* > 0, the mathematical form of the SIR model with birth and death is presented as (2.2).

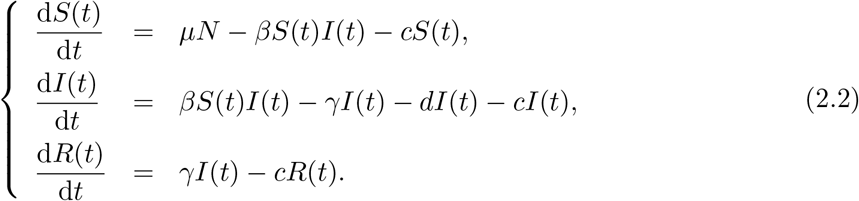

### 2.2 SEIR Model

In this section, we first introduce the classic SEIR model in Section 2.2.1. Then it comes to the SEIR model with death and migration in Section 2.2.2.

#### 2.2.1 Classic SEIR Model

In real life, many epidemics (such as influenza) have an incubation period, so we need to consider exposed people when constructing the corresponding epidemic dynamic models. The classic SEIR model proposed in [19] is suitable for this circumstance. The classic SEIR model divides the population into four groups: susceptible (*S*), exposed (*E*), infected (*I*), and recovered (*R*), and the numbers of these four groups at time *t* are denoted by *S*(*t*), *E*(*t*), *I*(*t*) and *R*(*t*), respectively. The flowchart of the classic SEIR model is presented as Figure 3, where *σ* is the rate of an exposed person turning into an infected person.

**Figure 3:**
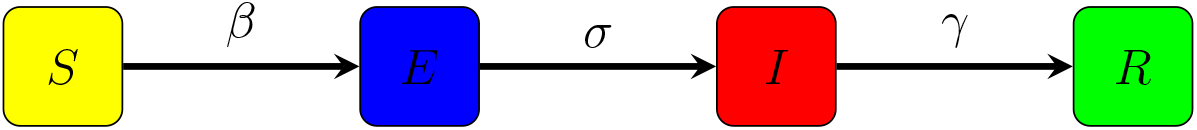
Flowchart of classic SEIR model

Based on the flowchart, the mathematical form of the classic SEIR model is defined as (2.3).

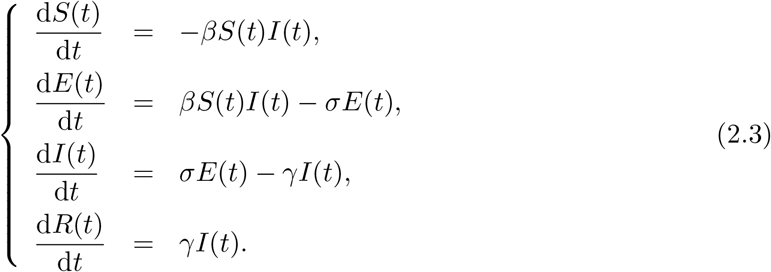

#### 2.2.2 SEIR Model with Death and Migration

In this section, we consider an SEIR model with death and migration. *A* donated as the number of migrations is a constant, and *d* and *c* are the same as Section 2.1.2. The flowchart of the SEIR model with death and migration is given in Figure 4.

**Figure 4:**
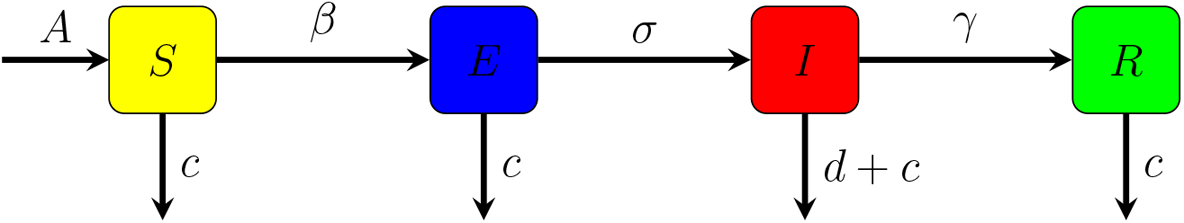
Flowchart of SEIR model with death and migration

According to the flowchart, the mathematical form of the SEIR model with death and migration is defined as (2.4).

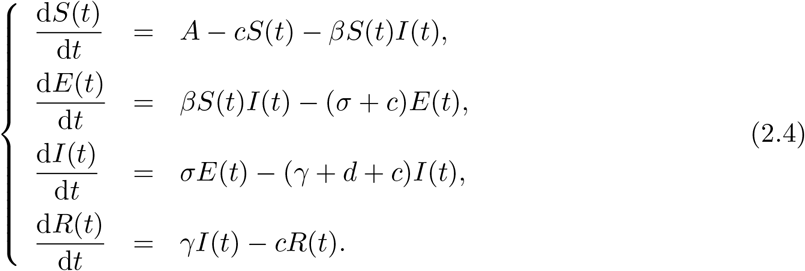

### 2.3 Basic Reproduction Number Theory

Regarding an epidemic, the problem we are most concerned about is whether the disease can spread among the population. The basic reproductive number proposed in [32] is an important indicator used to describe the ability of an epidemic to spread. Definition 2.1 provides the detailed definition of the basic reproduction number.

#### Definition 2.1

The basic reproduction number represents the number of susceptible persons infected with the disease by each infected person during the entire infectious period at the early stage of the epidemic, denoted as *R*_0_.

For a basic reproductive number, *R*_0_ = 1 is the threshold for judging whether an epidemic is prevalent. When *R*_0_ *<* 1, even if no prevention and control measures are taken, the number of people infected by the epidemic will decrease. On the contrary, when *R*_0_ > 1, the number of people infected will increase.

The basic reproductive number is calculated by the next generation matrix approach [32]. The detailed calculation steps are as follows.

Firstly, we divide the whole population into *n* compartments, let *x* = (*x*_1_, ⋯, *x*_*n*_)^T^, with each *x*_*i*_ *≥* 0, be the number of individuals in each compartment. It should be noted that only the new infected population is considered as *x*_*i*_, e.g., exposed (*E*) and infected (*I*). The epidemic dynamic model can be presented as follows:

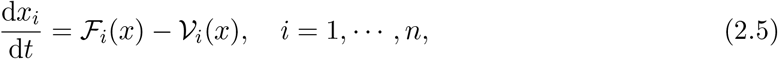

where 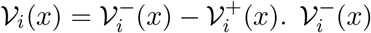 is the rate of transfer of individuals out of compartment 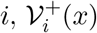 is the rate of transfer of individuals into compartment *i* by all other means, and ℱ_*i*_(*x*) is the rate of appearance of new infections in compartment *i*.

Secondly, calculate *F* and *V*, which defined as:

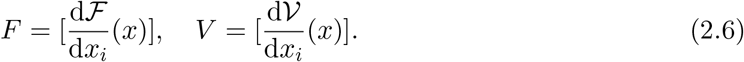

Thirdly, calculate the basic reproduction number *R*_0_, which is:

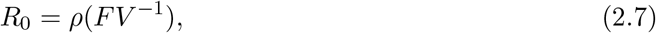

where *ρ*(*A*) denotes the spectral radius (maximum eigenvalue) of a matrix *A* and *A*^*−*1^ is the transpose of a matrix *A*.

## 3 Establishment of the SEIDR Model

It is widely known that COVID-19 has a significant incubation period, so the type SEIR model is suitable for this infectious disease. Based on the SEIR model, we add a compartment ‘diagnosed (*D*)’ into the model. The flowchart of the new SEIDR model is shown in Figure 5.

**Figure 5:**
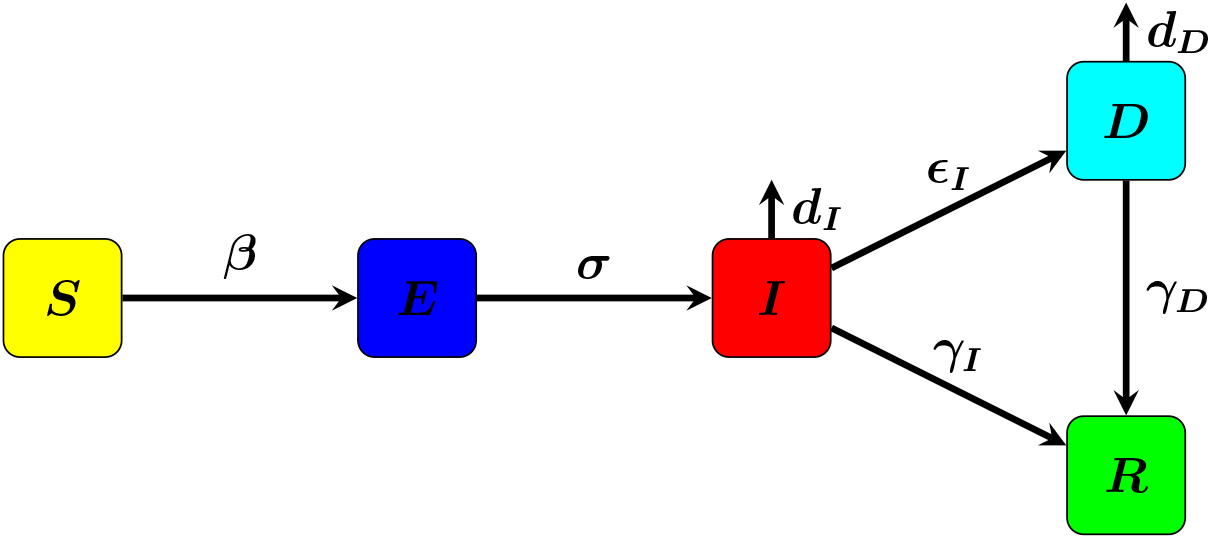
Flowchart of the SEIDR model

For the parameters, *β* is the contact rate, and 1*/σ* refers to the mean latency period of COVID-19. Once the virus infects a person, the average confirmed period is defined by 1/*ε*_*I*_. *γ*_*I*_ represents the self-cure rate, and *γ*_*D*_ is the recovery rate of hospitalized patients. For the fatality rate, we denote *d*_*D*_ and *d*_*I*_ to distinguish the death rate with and without medical treatment, respectively. The mathematical form of the SEIDR model is defined as (3.1).

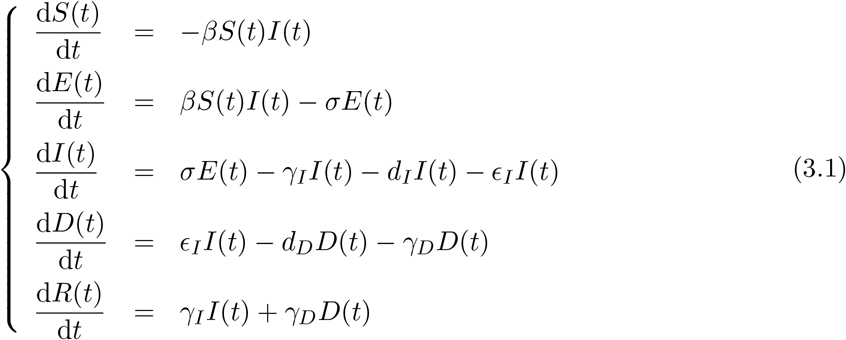

The basic reproduction number *R*_0_ is calculated based on the next generation matrix approach introduced in Section 2.3. According to the model, the newly infected population is considered, which means *x* = (*E, I*)^T^. *F* and *V* are calculated from (2.5).

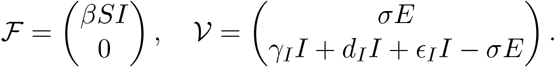

Then *F* and *V* are generated according to (2.6).

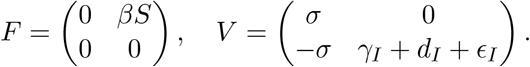

By solving the inverse of matrix *V, V* ^*−*1^ is received:

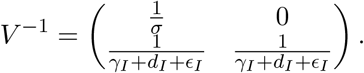

Based on (2.7), *FV* ^*−*1^ is needed:

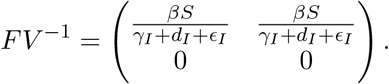

Since the spectral radius of a matrix *A* is the maximum eigenvalue of *A*, we define *X* = (*x*_1_, *x*_2_)^(T)^ and *λ* = (*λ*_1_, *λ*_2_)^(T)^, so that

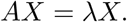

which means

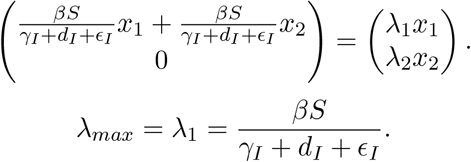

Thus, the basic reproduction number is:

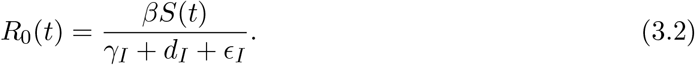

### 3.1 Solving Parameters

To solve (3.1), first, we need to solve the contact rate *β* and self-cure rate *γ*_*I*_ through the least square method. Let the two related variables be *x* and *y*, and their relationship can be expressed as *y* = *f* (*x*). There are parameters *α*_1_, *α*_2_, ⋯, *α*_*m*_ in function *f* (*x*). The observed dataset is (*x*_*i*_, *y*_*i*_), *i* = 1, 2, ⋯, *N*, and *N* is much larger than *m*. The error generated in each observation is

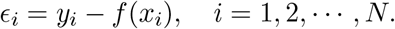

To control the error in every observed point, it is more appropriate to use the overall error, which defined as

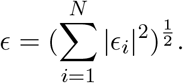

Thus, we seek for the parameters *α*_1_, *α*_2_, ⋯, *α*_*m*_ so that

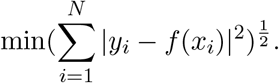

In practice, the population of diagnosed (*D*) is the only data we have. So the problem that needs to be solved is

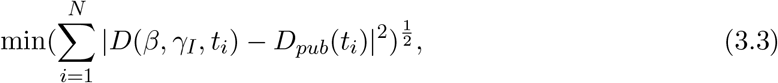

where *D*_*pub*_(*t*_*i*_) is the published population of diagnosed in day *t*_*i*_. To numerically solve (3.3), we choose the *fmincon* function in Matlab as the solver. It includes kinds of classic numerical solvers, including interior-point methods, trust region methods, quadratic sequence programs, etc. The *fmincon* function allows (3.3) to reach minimum and generate corresponding *β* and *γ*_*I*_.

### 3.2 Solving the Dynamic Model

When the contact rate *β* and self-cure rate *γ*_*I*_ are received, the SEIDR dynamic model needs to be solved. The Runge-Kutta method is a classic and efficient tool for numerically solving ordinary differential equations (ODEs). Assuming that the solution of an ODE *f* (*t, y*) is *y*, we need the initial values of the solution *y*_0_ and the total time *T*. The time *T* needs to be discreted equally into a time series {*t*_0_, *t*_1_, *t*_2_, ⋯, *t*_*N*_} and the time interval is *h*. If the solution *y* is smooth enough, it can be expressed as the Taylor expansion

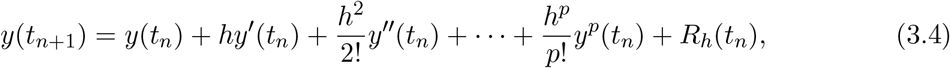

where

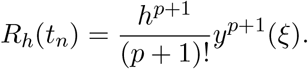

Since it would be too complex to calculate the higher derivatives in computers, the Runge-Kutta method uses the linear combination of *f* (*t, y*) to avoid the problem. The most commonly used method is the four stages Runge-Kutta scheme:

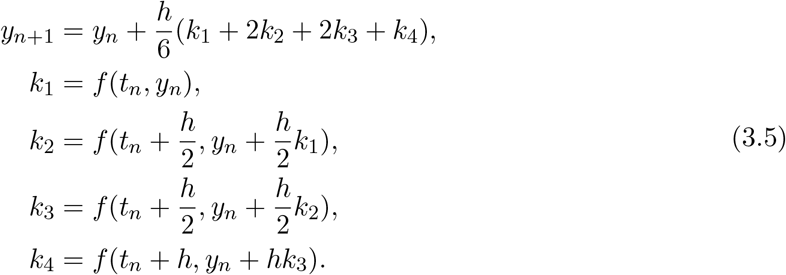

The error of (3.5) is

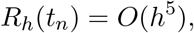

which means the fifth order infinitesimal quantity of *h*. To solve the SEIDR model, we need the initial values of the compartments *S*_0_, *E*_0_, *I*_0_, *D*_0_, and *R*_0_, which are inferred from actual data, and the total time *T*. Using the Runge-Kutta method, the population of every compartment every day can be calculated.

## 4 Numerical Experiments

In this section, we will use the SEIDR model to simulate the COVID-19 epidemics in Argentina, Indonesia, Mexico, and South Africa. The data on new infections daily is downloaded from World Health Organization (WHO). Since our model has not considered any prophylactic measures or vaccines, it is suitable for epidemics in the early stage. Specifically, the time range we focus on is Mar. 1st, 2020, to Jun. 1st, 2020. Predictions of new infections and the corresponding actual values are shown in Figure 6.

**Figure 6:**
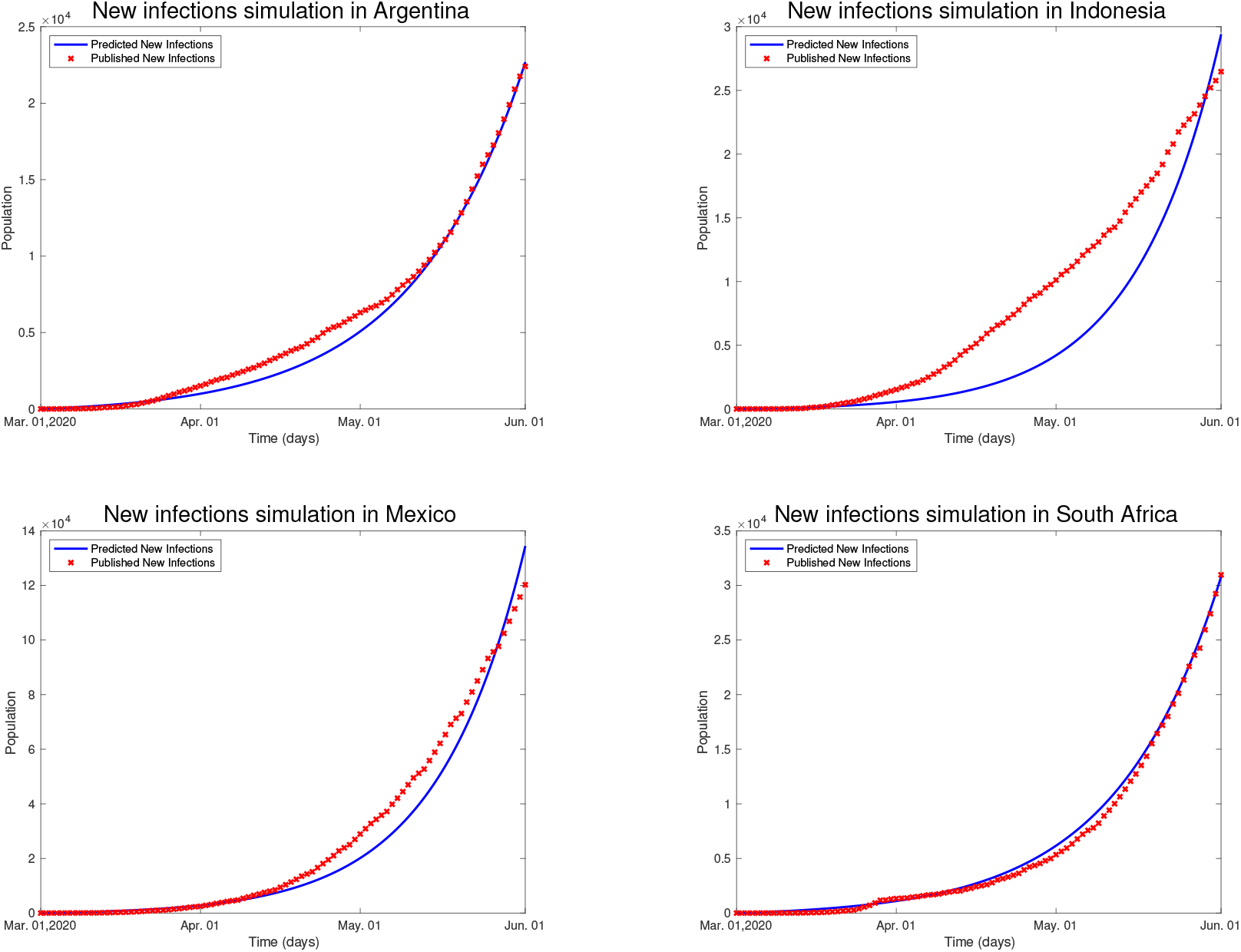
Simulations and real values on new infections

The simulation results show that our model fits reality well in most cases. However, if countries like Indonesia took severe prophylactic measures, the simulation may need to be revised. The results of the basic reproduction numbers are shown in Figure 7. The red dashed line is the baseline, and every country’s basic reproduction number is larger than 1. This indicates that COVID-19 spreads rapidly in every country, and the higher the basic reproduction number, the faster it spreads.

**Figure 7:**
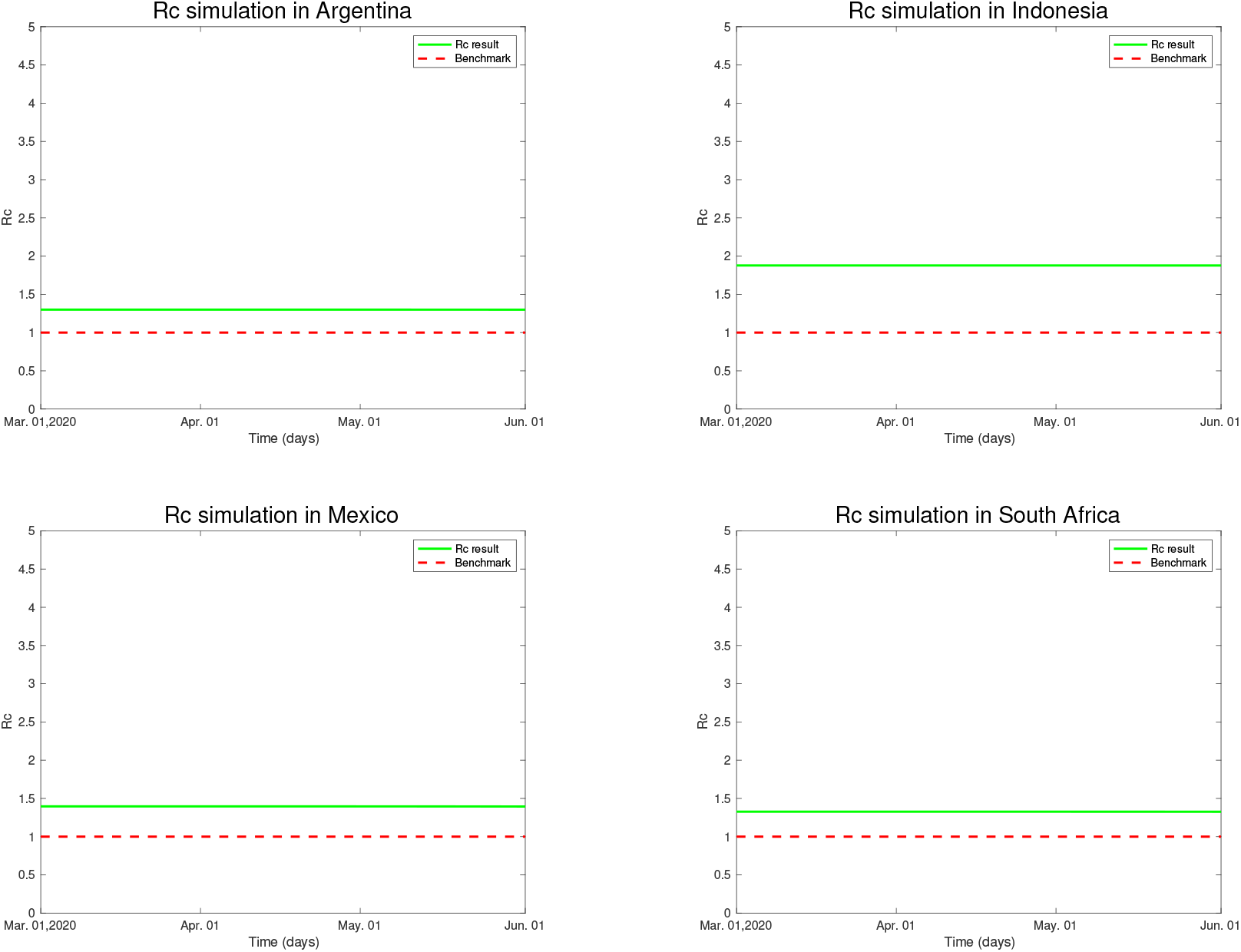
Results of the basic reproduction numbers

The initial parameters, such as *S*_0_, and the diseases’ parameters, such as recovery rates, are considered based on the countries’ situations. They are shown along with calculated parameters contact rate *β* and self-cure rate *γ*_*I*_ in Table 1.

**Table 1:** Parameters for Model

| Parameter | Argentina | Indonesia | Mexico | South Africa |
| --- | --- | --- | --- | --- |
| $\beta$ | 1.00e-09 | 1.00e-09 | 1.00e-09 | 7.46e-09 |
| $\sigma$ | 1/3 | 1/7 | 1/3 | 1/3 |
| $\gamma_I$ | 0.01 | 0.01 | 0.01 | 4.89e-12 |
| $\gamma_D$ | 0.01 | 0.01 | 0.01 | 5.38e-12 |
| $\epsilon_I$ | 1/3 | 1/5 | 1/3 | 1/3 |
| $d_D$ | 2.20e-06 | 2.60e-06 | 2.20e-06 | 1.08e-15 |
| $d_I$ | 3.30e-06 | 3.90e-06 | 3.30e-06 | 1.61e-15 |
| $S_0$ | 45377000 | 271000000 | 129000000 | 59309000 |
| $E_0$ | 100 | 100 | 200 | 100 |
| $I_0$ | 3 | 5 | 3 | 3 |
| $D_0$ | 1 | 1 | 1 | 1 |
| $R_0$ | 0 | 0 | 0 | 0 |

## 5 Conclusion

In this article, we first review the history of infectious diseases and COVID-19 and introduce the history of mathematical research on infectious diseases and the related achievements of mathematical analysis on COVID-19. Then we present some simple epidemic dynamics models - classic SIR model, SIR model with birth and death, classic SEIR model, and SEIR model with death and migration. At the same time, we introduce the basic reproductive number theory and its calculation method - the next generation matrix approach. Afterward, we propose the SEIDR model considering that only some infected persons are diagnosed, and introduce its basic reproduction number calculation process, the method of solving parameters, and the method of solving the model in detail. Finally, we use the model to simulate the COVID-19 epidemic in Argentina, Indonesia, Mexico, and South Africa. The results show that the SEIDR model can accurately simulate the early spread of the epidemic. However, the model still has shortcomings: for example, it does not consider some countries’ home isolation policies, nor does it consider the situation that some COVID-19-infected people have no apparent symptoms. Next, we will consider more complex situations and build corresponding models.

## Data Availability

All data produced in the present study are available upon reasonable request to the authors

